# Income-based inequalities in self-reported moderate-to-vigorous physical activity among adolescents in England and the United States: a cross-sectional study

**DOI:** 10.1101/2020.05.15.20102673

**Authors:** Shaun Scholes, Jennifer S Mindell

**Affiliations:** Research Department of Epidemiology and Public Health, UCL, United Kingdom

**Keywords:** moderate-to-vigorous physical activity, adolescents, England, United States, inequalities, hurdle models

## Abstract

**Objective:** Quantify inequalities in self-reported moderate-to-vigorous physical activity (MVPA) in England and the United States (US).

**Design:** Population-based cross-sectional study.

**Participants:** 4019 adolescents aged 11–15 years in England (Health Survey for England 2008, 2012, 2015) and 4312 aged 12–17 years in the US (National Health and Nutrition Examination Survey 2007–16).

**Main outcome measures:** Three aspects of MVPA: (1) doing any, (2) average min/day (MVPA: including those who did none), and (3) average min/day conditional on participation (MVPA-active). Using hurdle models, inequalities were quantified using the absolute difference in marginal means (average marginal effects: AMEs).

**Results:** In England, adolescents in high-income households were more likely than those in low-income households to have done any formal sports/exercise in the last seven days (boys: 11%; 95% CI: 4% to 17%; girls: 13%; 95% CI: 6% to 20%); girls in high-income households did more than their low-income counterparts (MVPA: 6 min/day, 95% CI: 2 to 9). Girls in low-income households spent more time in informal activities than girls in high-income households (MVPA: 21 min/day; 95% CI: 10 to 33), whilst boys in low-income versus high-income households spent longer in active travel (MVPA: 21 min/week; 95% CI: 8 to 34). In the US, in a typical week, recreational activity was greater among high-income versus low-income households (boys: 15 min/day; 95% CI: 6 to 24; girls: 19 min/day; 95% CI: 12 to 27). In contrast, adolescents in low-income versus high-income households were more likely to travel actively (boys: 11%; 95% CI: 3% to 19%; girls: 10%; 95% CI: 3% to 17%) and do more.

**Conclusions:** Policy actions and interventions are required to increase MVPA across all income groups in England and the US. Differences in formal sports/exercise (England) and recreational (US) activities suggest that additional efforts are required to reduce inequalities.

## INTRODUCTION

Being physically active benefits mental, physical and social health in a dose-response manner,[1] and is beneficial for higher academic achievement,[2] yet global data for 2016 show that more than 80% of school-going adolescents aged 11–17 years did not meet the World Health Organization’s (WHO) daily minimum recommendation of one hour of physical activity (PA).[3] Socioeconomic inequalities in adolescent PA is an additional national and international concern:[4] evidence suggests these are domain-specific, with levels of activity in sports especially higher among the most advantaged.[5]

Whilst enabling assessment against PA recommendations, grouping a quantitative variable such as the minutes-per-day (min/day) that adolescents typically spend engaged in moderate-to-vigorous intensity PA (MVPA) into a binary or ordinal variable loses information and weakens statistical power.[6] Yet analysing the quantitative variable is also problematic as MVPA distributions are not typically normally distributed but contain a stack of zeros (adolescents not doing any) and are positively skewed (high values for a small number who are highly active).[7] Such data can be transformed to meet normality assumptions [7] but findings based on a single-equation regression model cannot identify potentially different determinants for participation and duration.[8]

Hurdle models [9] can handle quantitative MVPA data that contains a stack of zeros and positive skewness, as separate models can be fitted for the binary outcome of participation and the quantitative outcome of the amount of time spent active (conditional on overcoming the “hurdle” of participation). Although popular in the economics literature,[10] no epidemiological studies to date have used hurdle models to quantify inequalities in adolescent MVPA. Yet such models could indicate, for example, whether adolescents living in high-income households are more likely to do any MVPA but, conditional on doing any, spend less time on average in MVPA than their counterparts in low-income households.[10]

Using nationally representative cross-sectional data for adolescents in England and the United States(US), we applied hurdle models to quantify and compare income-based inequalities in self-reported total and domain-specific MVPA. We hypothesised that adolescents in high-income versus low-income households have a higher propensity to do any, and that conditional on taking part, spend more time on average being active.

## METHODS

Our study makes comparisons in inequalities in self-reported MVPA among adolescents in England and the US using nationally representative cross-sectional data.

### Data sources and study populations

#### England

The Health Survey for England (HSE) is used to monitor progress on numerous national health objectives, including PA for younger (aged 2–4 years) and older (5–15 years) children.[11–13] Details of sample design and data collection are described elsewhere.[14] Briefly, new, nationally-representative samples of people living in private households are drawn annually using multistage stratified probability sampling. We used the most recent surveys (2008, 2012, 2015) that included questions on children’s PA.[11–13] Up to two children aged 0–15 years were selected at each household in 2008 and 2012; a limit of four was used in 2015 (maximum two aged 13–15 years, interviewed directly, and maximum two aged 0–12 years, where a parent/guardian provided the information). Interviewers measured participants’ height and weight and assessed demographics and health behaviours including PA. The household response rate ranged from 64% in 2008 to 60% in 2015.

We restricted the analytical population in this study to adolescents aged 11–15 years; participants aged 16 years or older are treated as adults, and so responded to a different PA questionnaire. Participants gave verbal consent for interview. Relevant national committees granted research ethics approval prior to the surveys.

#### United States

The National Health and Nutrition Examination Survey (NHANES) uses a complex, stratified, multistage probability cluster sampling design. Details on sample design and data collection are described elsewhere.[15] Briefly, data collection is based on a nationally representative sample covering all ages of the civilian noninstitutionalised population. During 2011–14, non-Hispanic Black, non-Hispanic Asian, and Hispanic persons, among other groups, were oversampled. All eligible members within a household were listed and a subsample of persons was selected based on gender, age, race/ethnicity, and income.[15] NHANES protocols were approved by the National Center for Health Statistics Ethics Review Board. Written informed consent was obtained from all participants before participation.

To allow broad comparisons with the HSE and WHO data,[3] we restricted the analytical population for this study to adolescents aged 12–17 years (less detailed questions are asked of 2–11 year-olds via a parental proxy). As the same PA questionnaire was used, we pooled five two-year cycles (2007–08, 2009–10, 2011–12, 2013–14, 2015–16). Overall, 4705 adolescents aged 12–17 had valid PA data. Of these, 393 had missing income data and were excluded from our complete-case analysis, leaving an analytical sample of 4312 adolescents.

### Data collection and derivation of PA outcomes

#### Health Survey for England

##### Formal and informal activities

Adolescents (or their parents/guardians: hereafter referred to as participants) were asked questions about PA conducted outside school hours in the seven days prior to the day of interview.

Participants were presented with two lists of physical activities: (i) formal activities: ten specific (e.g. individual and team sports/exercise such as football, workout with gym machines) plus up to five ‘other’ activities; and (ii) informal activities: nine specific activities (e.g. cycling excluding to/from school; walking excluding to/from school; active play). For each activity identified, participants were asked to recall on which days they took part; and on each day, how long they spent engaged in that activity (with no specified minimum duration). Each activity was assumed to be at least moderately-intensive.

##### Active travel

Participants who had been to school on at least one day in the seven days prior to interview were asked whether they had walked or cycled all or part of the way to or from school on any of those days (positive responses: walking, cycling, or both). If the participants had walked, they were asked: (i) the number of days they walked to school, (ii) the number of days they walked from school, and (iii) how long it usually takes to walk to school (an average was given if the journeys to and from school differed). These questions were repeated for cycling. Each activity was assumed to be at least moderately-intensive.

##### Derivation of outcomes

Outcomes were domain-specific: formal activities; informal activities; and active travel. Due to the difference in questionnaire format (daily assessment for formal- and informal-activities; weekly for active travel), total MVPA was calculated as the sum of formal and informal activities only. This was truncated at 40 hours/week to minimise unrealistic values. Weekly totals (expressed in minutes) were divided by seven to calculate min/day. Time spent in active travel was obtained by multiplying the number of journeys (to and/or from school) by the usual time spent travelling (expressed as min/week). Those who had not attended school were included in all analyses but were allocated zero time for active travel.[16]

##### Socioeconomic position and confounders

Household income was our chosen marker of socioeconomic position (SEP). The household reference person reports annual gross household income via a showcard (31 bands ranging from ‘less than £520’ to ‘£150 000+’). Household income was equivalised (McClements scale[17]), and grouped into tertiles (lowest, middle, highest). Body mass index (BMI) was calculated from valid weight and height measurements as weight in kilogrammes divided by height in metres squared. Three weight status categories were derived based on age (categorised in six-month bands) and the gender-specific UK National BMI centiles classification:[18] healthy weight (a BMI-for-age below the 85^th^ percentile), overweight (85^th^ to below the 95^th^ percentile), and obese (≥ 95^th^ percentile).

#### National Health And Nutrition Examination Survey

##### Global Physical Activity Questionnaire

An adapted version of the Global Physical Activity Questionnaire (GPAQ), developed by the WHO,[19] was administered directly to 12–15 year-olds at the Mobile Examination Center (MEC), and during in-home interviews to 16–17 year-olds.[20] The GPAQ captures aerobic PA in three domains: recreational, active transportation, and work (e.g. paid or unpaid work, household chores, yard work). For the recreational- and work-domains, participants are asked whether they do any vigorous-intensity activities (VPA) that “cause large increases in breathing or heart rate for at least 10 minutes continuously” in a typical week; those answering positively, are asked on how many days in a typical week they do VPA, and for how much time they spend doing VPA on a typical day. Similar questions were asked for moderate-intensity activities: those that “cause a small increase in breathing or heart rate”. For active transportation, participants are asked whether they walk or use a bicycle for at least 10 minutes continuously to get to and from places; those answering positively, are asked on how many days in a typical week they engage in such travel, and for how much time they spend travelling actively on a typical day (walking and bicycling are not assessed separately).

Outcomes were truncated at 40 hours/week to minimise unrealistic values. Total MVPA was calculated as the sum across the three domains. Frequency (number of days/week) and duration (average min/day) were multiplied and then divided by seven to calculate min/day MVPA for total and domain-specific MVPA.[7]

##### Socioeconomic position and confounders

Household income was reported by the household reference person. The inflation-adjusted family income-to-poverty ratio (FIPR) is calculated by dividing family income by a poverty measure specific for family size. Larger FIPRs indicate higher income and was categorized as in other studies [21,22] as low (<1.3), middle (>1.3 to 3.5), and high (>3.5) (high-income). Race/ethnicity was categorised as non-Hispanic White, non-Hispanic Black, Mexican-American, and other. Three weight status categories were based on the Center for Disease Control and Prevention’s (CDC) gender-specific 2000 BMI-for-age growth charts for the US:[22] healthy weight, overweight, and obese were defined analogously to that described above for the HSE.

#### Statistical analysis

##### Sample characteristics

Data was pooled over the survey years to increase precision. Differences in age, race/ethnicity (US), and weight status were estimated by income, using Rao-Scott tests for independence.[23] To address potential bias in the composition of the analytical sample, HSE analyses were weighted using the appropriate selection and non-response weight; NHANES analyses used the combined two-year MEC sample weights which account for differential probabilities of selection, non-response, and differences between the final sample and the US civilian non-institutionalised population.[15]

##### Hurdle models

To handle quantitative MVPA data that contains a stack of zeros and positive skew, we used the Cragg hurdle model,[9] which comprises two parts: a selection/participation model and a latent model. The former is used to examine differences in the propensity for the quantitative outcome to take positive values versus zero, whilst the latter examines differences in the positive, non-zero part of the distribution among those with non-zero values. Reflecting the difference in questionnaires, the lowest (observed) value for positive MVPA was five minutes (i.e. 0.071 min/day) in HSE and ten minutes (i.e. 1.43 min/day) in NHANES. In our analyses, the selection model assessed the influence of household income status on the binary outcome of participation (any versus none), whilst the latent model assessed its influence on the amount of time spent active, conditional on doing any MVPA (hereafter referred to as MVPA-active). We specified a probit model for the former and an exponential form for the latter. Each model contained income (as a three-category variable) and the confounders listed above.

Based on the model estimates, three sets of marginal means by income were calculated, evaluated at fixed values of the confounders. These sets correspond to different definitions of the expected value of MVPA:[24] (i) the probability of doing any, (ii) the average min/day MVPA for all participants(the unconditional mean), including those who did none; and (iii) the average min/day MVPA conditional on participation (MVPA-active). Average marginal effects (AMEs), representing inequalities after confounder adjustment, were quantified by computing the absolute difference in the marginal means (low-income households as reference).

We decided, a-priori, to conduct gender-stratified analyses due to expected differences in MVPA levels and inequalities in these as reported in the literature.[20,25] Dataset preparation and analysis was performed in SPSS V22.0 (SPSS IBM Inc., Chicago, Illinois, USA) and Stata V15.0 (College Station, Texas, USA) for the HSE; datasets are available via the UK Data Service (http://www.ukdataservice.ac.uk).[26–28](http://www.ukdataservice.ac.uk).[26–28] Stata was used to prepare and analyse NHANES; datasets are available via the CDC website (https://www.cdc.gov/nchs/nhanes). All analyses were performed using the survey procedures to account for the complex survey designs, including the geographical clustering of participants in primary sampling units. Two-sided P-values < 0.05 were considered statistically significant. This manuscript was written according to the Strengthening the Reporting of Observational Studies in Epidemiology statement.

### Patient and public involvement

Patients or the public were not involved in the design, or conduct, or reporting, or dissemination plans of our research (which involves secondary analysis of existing data).

## RESULTS

### Analytical samples

In England, 4897 adolescents aged 11–15 years participated in one of the three surveys (2008, 2012, 2015), of whom 4874 had valid PA data. Of these, 855 had missing income data and were excluded from our complete-case analysis, leaving an analytical sample of 4019 adolescents. In the United States, 4705 adolescents aged 12–17 had valid PA data. Of these, 393 had missing income data and were excluded from our complete-case analysis, leaving an analytical sample of 4312 adolescents.

### Sample characteristics

Information on key demographics by household income status is presented in Table 1. Adolescents in high-income households in the US were predominantly non-Hispanic White, whilst the proportions with healthy weight were highest in high-income households among both genders in both countries.

**Table 1:** Key variables by income tertile and gender, Health Survey for England (HSE: 2008, 2012, 2015) and National Health and Nutrition Examination Survey (NHANES 2007–2016)

|  | Boys |  |  |  |  | Girls |  |  |  |  |
| --- | --- | --- | --- | --- | --- | --- | --- | --- | --- | --- |
|  | Income |  |  |  |  | Income |  |  |  |  |
|  | All | Lowest | Middle | Highest | P-value<sup>a</sup> | All | Lowest | Middle | Highest | P-value<sup>a</sup> |
|  | N (%) | N (%) | N (%) | N (%) |  | N (%) | N (%) | N (%) | N (%) |  |
| **England:** |  |  |  |  |  |  |  |  |  |  |
| **All** | 2387 (100) | 714 (100) | 725 (100) | 529 (100) | - | 2487 (100) | 759 (100) | 701 (100) | 591 (100) | - |
| **Age-group:** |  |  |  |  |  |  |  |  |  |  |
| 11-12 | 988 (41) | 303 (42) | 311 (42) | 217 (39) | 0.506 | 1040 (41) | 308 (40) | 309 (42) | 259 (44) | 0.915 |
| 13-15 | 1399 (59) | 411 (58) | 414 (58) | 312 (61) |  | 1447 (59) | 451 (60) | 392 (58) | 332 (56) |  |
| **Weight status:<sup>b</sup>** |  |  |  |  |  |  |  |  |  |  |
| Healthy weight | 1346 (57) | 373 (53) | 418 (58) | 335 (65) | 0.033 | 1417 (58) | 401 (54) | 390 (57) | 385 (66) | <0.001 |
| Overweight | 340 (14) | 103 (14) | 105 (14) | 82 (15) |  | 357 (14) | 97 (12) | 116 (16) | 86 (15) |  |
| Obese | 415 (17) | 138 (19) | 122 (17) | 75 (13) |  | 378 (15) | 137 (18) | 114 (16) | 64 (10) |  |
| Missing | 286 (12) | 100 (14) | 80 (12) | 37 (7) |  | 335 (14) | 124 (16) | 81 (11) | 56 (9) |  |
| **United States:** |  |  |  |  |  |  |  |  |  |  |
| **All** | 2431 (100) | 917 (100) | 796 (100) | 509 (100) | - | 2274 (100) | 862 (100) | 775 (100) | 453 (100) | - |
| **Age-group:** |  |  |  |  |  |  |  |  |  |  |
| 12-15 | 1595 (67) | 619 (67) | 519 (66) | 334 (68) | 0.738 | 1470 (65) | 560 (64) | 508 (67) | 291 (63) | 0.381 |
| 16-17 | 836 (33) | 298 (33) | 277 (34) | 175 (32) |  | 804 (35) | 302 (36) | 267 (33) | 162 (37) |  |
| **Race/ethnicity:** |  |  |  |  |  |  |  |  |  |  |
| Non-Hispanic White | 689 (57) | 192 (34) | 223 (56) | 246 (78) | <0.001 | 617 (59) | 178 (38) | 217 (61) | 201 (77) | <0.001 |
| Non-Hispanic Black | 618 (14) | 263 (22) | 219 (15) | 87 (6) |  | 553 (14) | 224 (20) | 192 (14) | 88 (8) |  |
| Mexican-American | 536 (13) | 250 (24) | 177 (14) | 45 (4) |  | 535 (13) | 264 (24) | 173 (12) | 38 (4) |  |
| Other | 588 (15) | 212 (20) | 177 (15) | 131 (12) |  | 569 (14) | 196 (19) | 193 (14) | 126 (11) |  |
| **Weight status:<sup>b</sup>** |  |  |  |  |  |  |  |  |  |  |
| Healthy weight | 1422 (63) | 515 (58) | 461 (62) | 327 (67) | 0.033 | 1306 (63) | 455 (54) | 427 (59) | 320 (75) | <0.001 |
| Overweight | 388 (16) | 142 (16) | 128 (17) | 72 (15) |  | 409 (17) | 165 (19) | 148 (18) | 64 (14) |  |
| Obese | 518 (21) | 219 (26) | 176 (20) | 84 (18) |  | 486 (20) | 212 (27) | 177 (23) | 56 (11) |  |

Figures are column percentages. Sample sizes are unweighted; percentages are weighted (not age-standardised).

### MVPA distributions

Boys and girls in England spent 96 and 70 min/day on average in total MVPA in the last seven days, respectively; equivalent figures for total MVPA in the US were 100 and 67 min/day. However, each distribution showed a stack of zeros (highest among girls in the US) and was positively skewed(Figure 1–2).

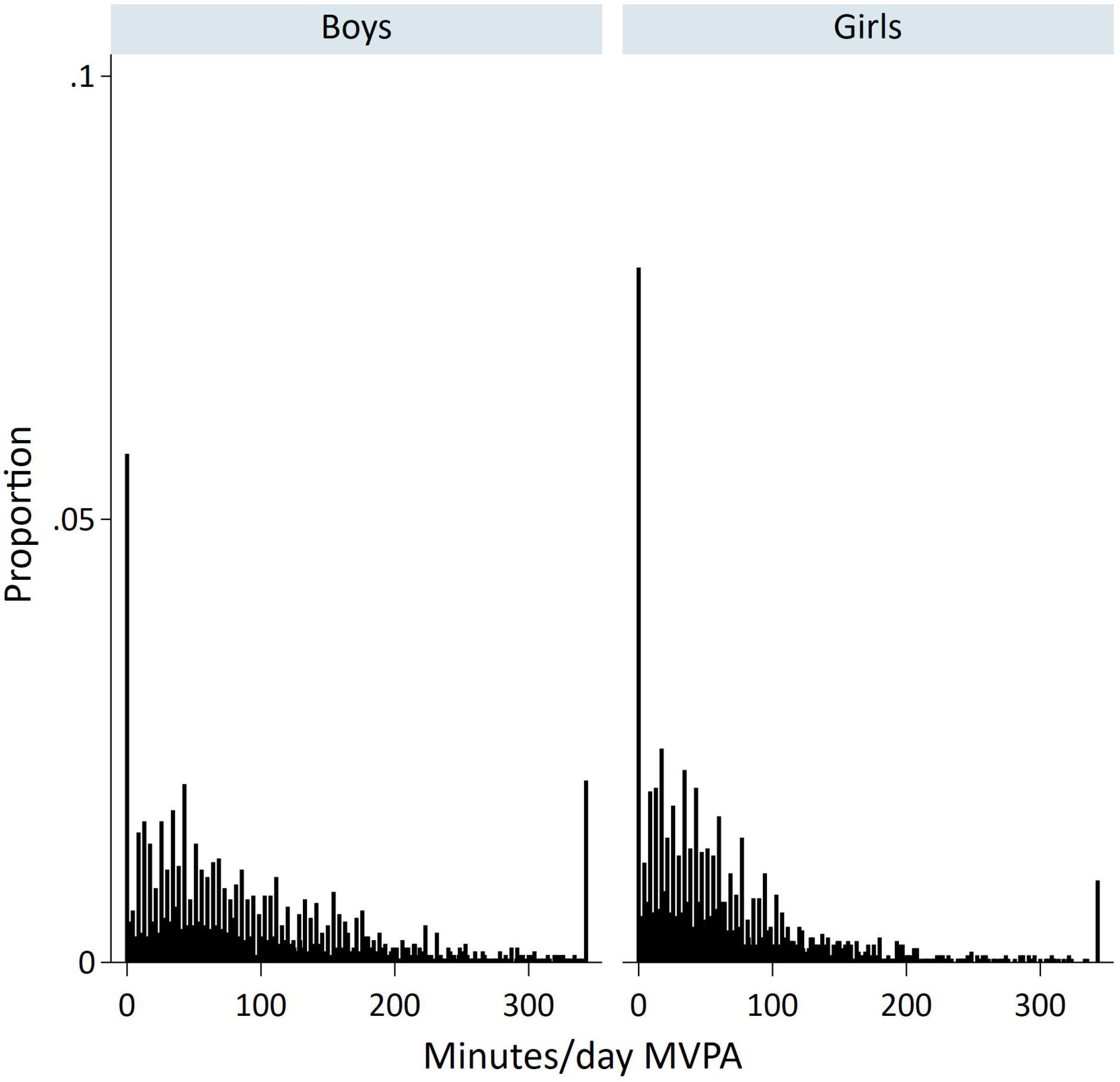
Figure 1.

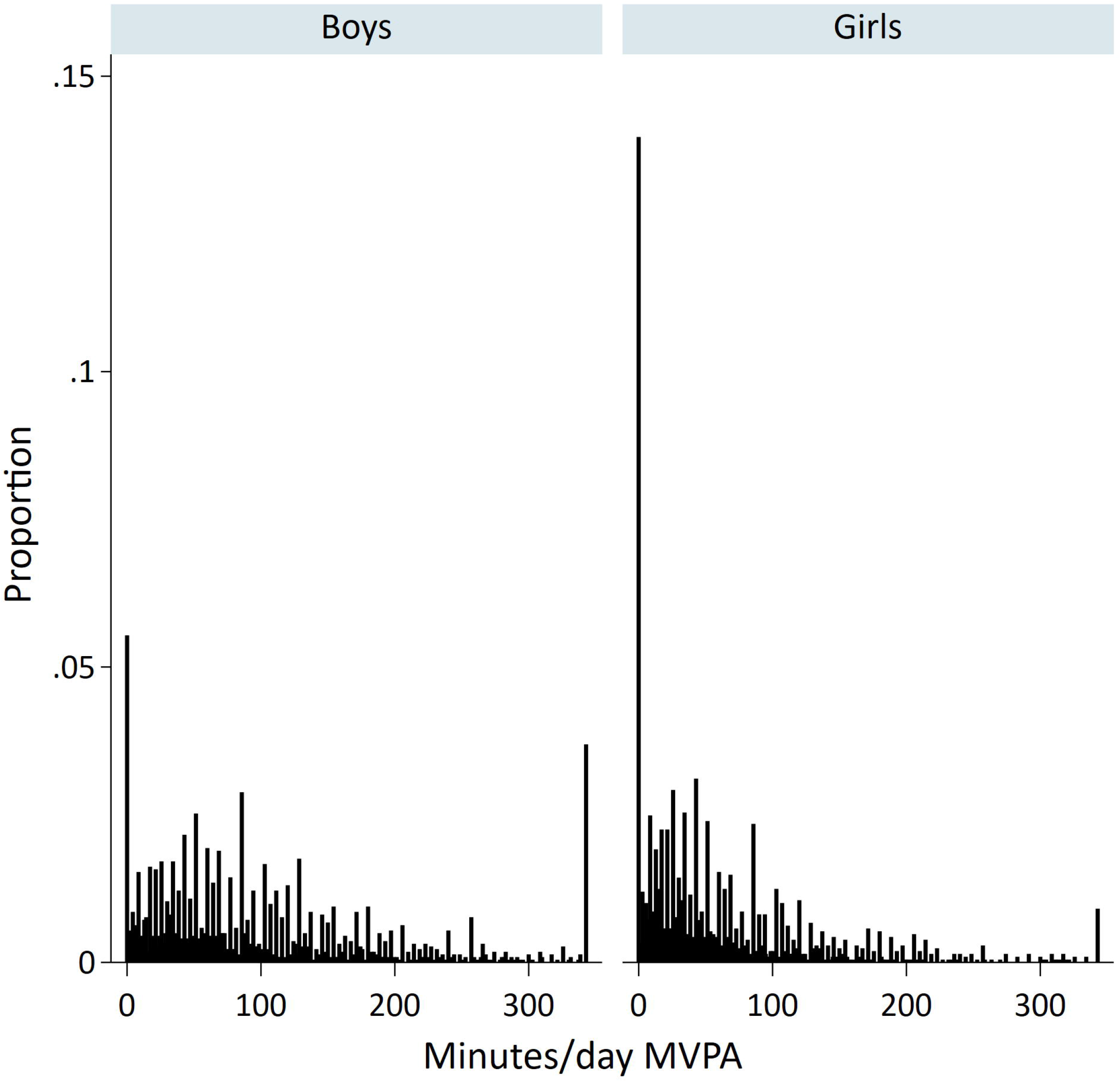
Figure 2.

### Hurdle models

Table 2 (England) and Table 3 (US) show the average marginal effects (AMEs) from the estimated hurdle models corresponding to the absolute difference in the income-specific marginal means for the binary outcome of participation (doing any versus none), and the quantitative outcomes of MVPA (including those who did none) and MVPA-active (conditional on those who did any). AMEs are shown graphically in Figure 3 (England) and Figure 4 (US).

**Table 2.** Parameter estimates from multivariable hurdle models (any participation and amount of time spent active), Health Survey for England 2008, 2012 and 2015.

|  | Any (%) |  | Unconditional: Mean MVPA<br>min/day |  | Conditional:<br>Mean MVPA-active min/day |  |
| --- | --- | --- | --- | --- | --- | --- |
|  | AME (95% CI)<sup>a</sup> | P-value | AME (95% CI)<sup>a</sup> | P-value | AME (95% CI)<sup>a</sup> | P-value |
| **Boys** |  |  |  |  |  |  |
| **Total:** |  |  |  |  |  |  |
| Middle vs lowest | 3 (0, 5) | 0.022 | 6.3 (-10.2, 22.8) | 0.454 | 3.0 (-14.1, 20.1) | 0.731 |
| Highest vs lowest | -1 (-4, 2) | 0.592 | 0.9 (-14.0, 15.7) | 0.910 | 2.0 (-13.1, 17.2) | 0.792 |
| **Formal activities:** |  |  |  |  |  |  |
| Middle vs lowest | 7 (1, 13) | 0.030 | 2.2 (-1.8, 6.3) | 0.284 | -0.3 (-6.0, 5.3) | 0.910 |
| Highest vs lowest | 11 (4, 17) | 0.001 | 2.3 (-1.7, 6.2) | 0.258 | -2.4 (-7.7, 2.9) | 0.367 |
| **Informal activities:** |  |  |  |  |  |  |
| Middle vs lowest | 4 (1, 8) | 0.004 | -0.8 (-16.6, 15.0) | 0.919 | -5.8 (-22.6, 11.0) | 0.501 |
| Highest vs lowest | -3 (-7, 1) | 0.207 | -8.4 (-22.7, 6.0) | 0.252 | -6.4 (-21.7, 8.9) | 0.410 |
| **Active travel:** |  |  |  |  |  |  |
| Middle vs lowest | 1 (-6, 7)<sup>b</sup> | 0.854 | 2.2 (-10.4, 14.7)<sup>b</sup> | 0.732 | 2.4 (-13.5, 18.3)<sup>b</sup> | 0.766 |
| Highest vs lowest | -8 (-15, -1)<sup>b</sup> | 0.022 | -20.6 (-33.5, -7.7)<sup>b</sup> | 0.002 | -20.3 (-38.4, -2.2)<sup>b</sup> | 0.028 |
| **Girls** |  |  |  |  |  |  |
| **Total:** |  |  |  |  |  |  |
| Middle vs lowest | -2 (-5, 0) | 0.084 | -5.4 (-19.6, 8.8) | 0.454 | -3.1 (-17.7, 11.6) | 0.682 |
| Highest vs lowest | 0 (-2, 3) | 0.958 | -5.1 (-18.5, 8.3) | 0.454 | -5.4 (-19.1, 8.2) | 0.437 |
| **Formal activities:** |  |  |  |  |  |  |
| Middle vs lowest | 4 (-2, 10) | 0.218 | 1.5 (-1.3, 4.4) | 0.293 | 1.1 (-3.7, 5.9) | 0.658 |
| Highest vs lowest | 13 (6, 20) | <0.001 | 5.5 (2.2, 8.8) | 0.001 | 3.8 (-1.3, 8.9) | 0.141 |
| **Informal activities:** |  |  |  |  |  |  |
| Middle vs lowest | -3 (-6, 1) | 0.147 | -11.6 (-24.4, 1.1) | 0.074 | -10.3 (-23.7, 3.1) | 0.132 |
| Highest vs lowest | -2 (-6, 1) | 0.198 | -21.4 (-32.9, -9.8) | <0.001 | -21.3 (-33.4, -9.2) | 0.001 |
| **Active travel:** |  |  |  |  |  |  |
| Middle vs lowest | 4 (-2, 10)<sup>b</sup> | 0.225 | 13.5 (-0.1, 27.2)<sup>b</sup> | 0.052 | 15.0 (-3.4, 33.3)<sup>b</sup> | 0.110 |
| Highest vs lowest | 0 (-7, 7)<sup>b</sup> | 0.969 | -3.0 (-15.4, 9.5)<sup>b</sup> | 0.637 | -5.9 (-22.9, 11.2)<sup>b</sup> | 0.501 |

<sup>a</sup> Adjusting for age and weight status. Missing weight status as additional category. AMEs evaluated at fixed values of the confounders: for adolescents aged 11-12 years and having a healthy weight (a BMI-for-age below the 85<sup>th</sup> percentile).
<sup>b</sup> Estimates for active travel are min/week.
AME: average marginal effect.

**Table 3.** Parameter estimates from multivariable hurdle models (any participation and amount of time spent active), NHANES 2007–2016.

|  | Any (%) |  | Unconditional: Mean MVPA<br>min/day |  | Conditional:<br>Mean MVPA-active min/day |  |
| --- | --- | --- | --- | --- | --- | --- |
|  | AME (95% CI)<sup>a</sup> | P-value | AME (95% CI)<sup>a</sup> | P-value | AME (95% CI)<sup>a</sup> | P-value |
| **Boys** |  |  |  |  |  |  |
| **Total:** |  |  |  |  |  |  |
| Middle vs lowest | 1 (-2, 3) | 0.474 | 3.6 (-10.2, 17.4) | 0.606 | 2.7 (-10.9, 16.4) | 0.693 |
| Highest vs lowest | 3 (0, 6) | 0.023 | 9.8 (-7.2, 26.8) | 0.254 | 6.4 (-10.3, 23.2) | 0.449 |
| **Recreational:** |  |  |  |  |  |  |
| Middle vs lowest | 3 (-1, 7) | 0.148 | 10.2 (2.8, 17.6) | 0.007 | 9.1 (1.2, 17.0) | 0.024 |
| Highest vs lowest | 6 (2, 10) | 0.005 | 15.1 (5.8, 24.3) | 0.002 | 11.8 (2.2, 21.4) | 0.016 |
| **Work:** |  |  |  |  |  |  |
| Middle vs lowest | 3 (-3, 9) | 0.304 | -1.7 (-9.2, 5.9) | 0.657 | -6.5 (-19.8, 6.9) | 0.339 |
| Highest vs lowest | 9 (2, 16) | 0.012 | -1.1 (-10.1, 7.9) | 0.808 | -10.7 (-25.4, 3.9) | 0.149 |
| **Active transport:** |  |  |  |  |  |  |
| Middle vs lowest | -9 (-14, -5) | <0.001 | -5.3 (-9.1, -1.5) | 0.007 | -4.5 (-10.1, 1.0) | 0.108 |
| Highest vs lowest | -11 (-19, -3) | 0.005 | -7.1 (-11.6, -2.7) | 0.002 | -7.7 (-14.3, -1.1) | 0.024 |
| **Girls** |  |  |  |  |  |  |
| **Total:** |  |  |  |  |  |  |
| Middle vs lowest | 2 (0, 5) | 0.061 | 11.3 (1.6, 21.0) | 0.022 | 10.0 (-0.1, 20.1) | 0.052 |
| Highest vs lowest | 4 (0, 7) | 0.034 | 15.8 (5.1, 26.4) | 0.004 | 13.7 (2.7, 24.7) | 0.015 |
| **Recreational:** |  |  |  |  |  |  |
| Middle vs lowest | 6 (2, 10) | 0.005 | 11.4 (3.9, 18.9) | 0.003 | 9.6 (1.1, 18.1) | 0.027 |
| Highest vs lowest | 10 (5, 16) | <0.001 | 19.5 (11.8, 27.1) | <0.001 | 15.7 (7.7, 23.6) | <0.001 |
| **Work:** |  |  |  |  |  |  |
| Middle vs lowest | 3 (-3, 8) | 0.351 | 1.6 (-3.0, 6.2) | 0.499 | 1.4 (-8.4, 11.1) | 0.778 |
| Highest vs lowest | 6 (-1, 13) | 0.077 | 0.9 (-3.8, 5.7) | 0.700 | -3.4 (-13.1, 6.2) | 0.478 |
| **Active transport:** |  |  |  |  |  |  |
| Middle vs lowest | -2 (-8, 5) | 0.624 | -2.4 (-5.3, 0.5) | 0.102 | -4.6 (-9.9, 0.7) | 0.085 |
| Highest vs lowest | -10 (-17, -3) | 0.007 | -6.7 (-9.5, -3.9) | <0.001 | -11.4 (-16.3, -6.6) | <0.001 |

<sup>a</sup> Adjusting for age, race/ethnicity and weight status. AMEs evaluated at fixed values of the confounders: for adolescents aged 12-15 years, non-Hispanic White, and having a healthy weight (a BMI-for-age below the 85<sup>th</sup> percentile). AME: average marginal effect.

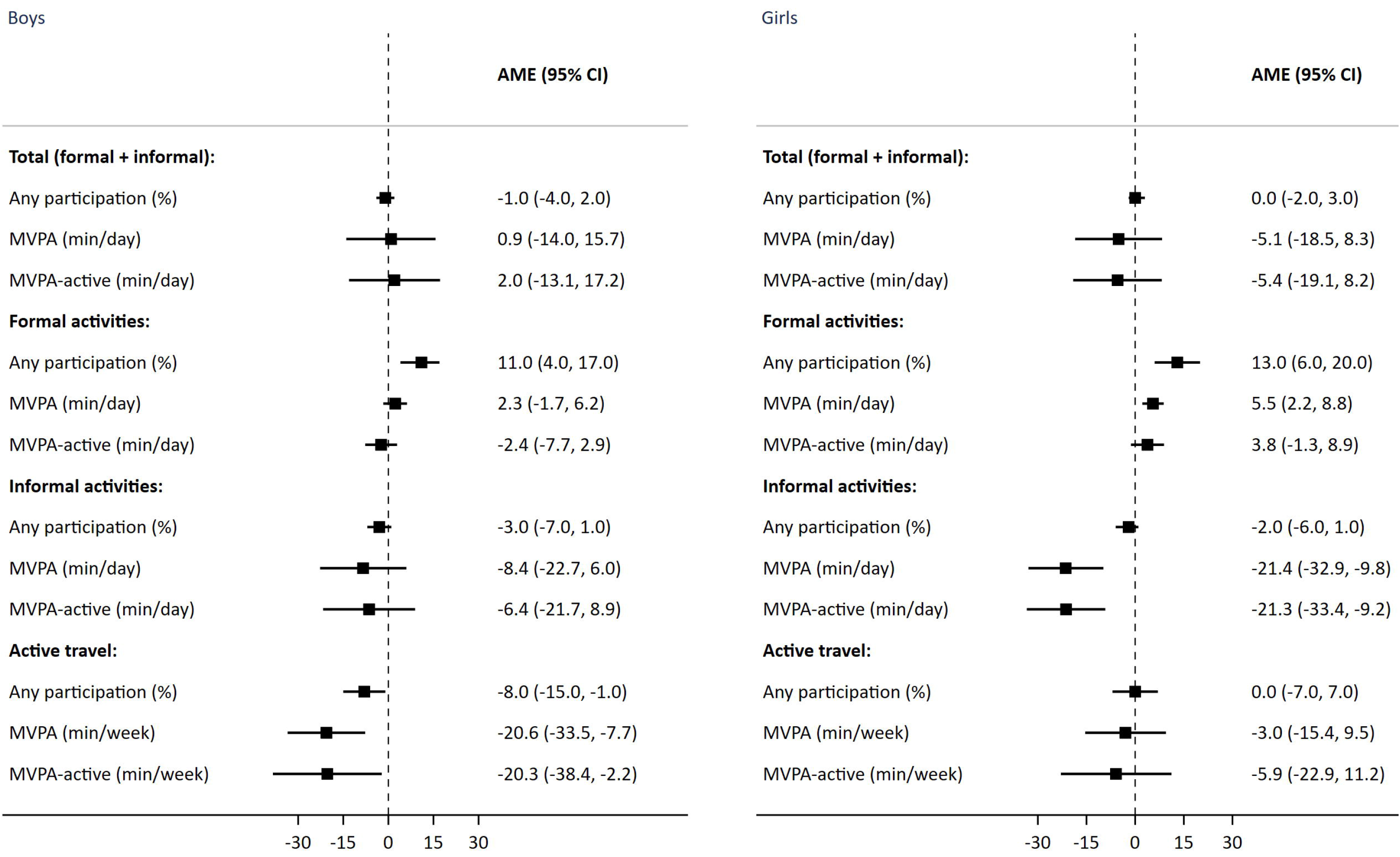
Figure 3

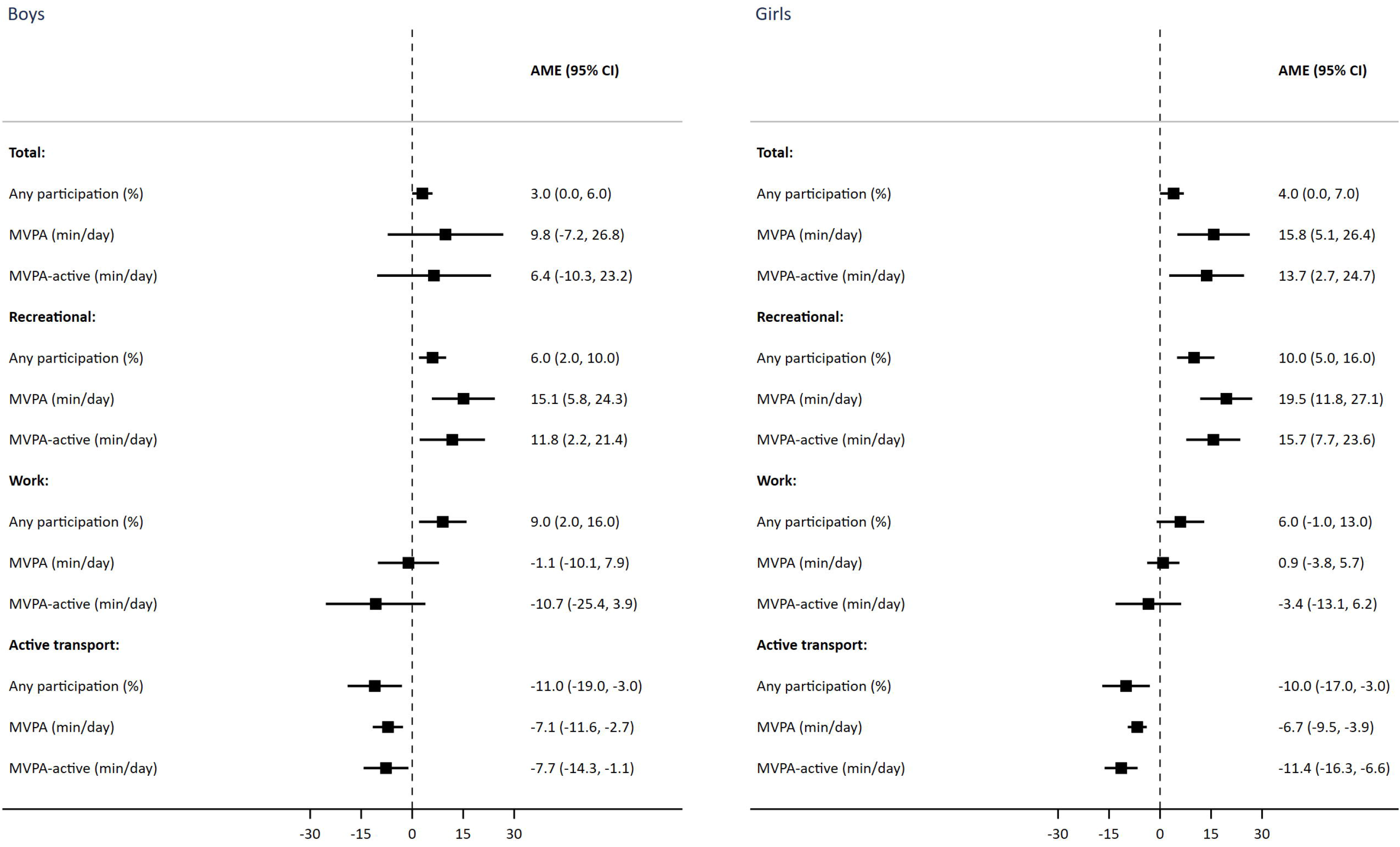
Figure 4

### Inequalities in MVPA in England

Among both genders, each of the three outcomes for total (i.e. formal and informal) MVPA showed similarities by income after confounder adjustment. However, this finding masked differences by gender, domain and outcome.

First, adolescents in high-income versus low-income households were more likely to have done any formal sports/exercise activity in the last seven days (AMEs boys: 11%; 95% CI: 4% to 17%; girls: 13%; 95% CI: 6% to 20%); whilst girls in low-income households spent more time being active than girls in high-income households did (AME formal MVPA: 6 min/day, 95% CI: 2 to 9). Secondly, girls in low-income households spent more time in informal activities than their counterparts in high-income households (informal MVPA: 21 min/day; 95% CI: 10 to 33; informal MVPA-active: 21 min/day; 95% CI: 9 to 33), whilst the differences in informal activities among boys were attenuated to the null. Thirdly, higher levels of active travel among boys in low-income versus high-income households were found for each of the three outcomes. The difference between boys in low-income versus high-income households in the probability of having done any active travel in the last seven days was 8% (95% CI: 1% to 15%). Among those who did any, boys in low-income versus high-income households spent 20 min/week more on average travelling actively (95% CI: 2 to 38).

### Inequalities in MVPA in the US

Among both genders, adolescents in high-income versus low-income households were more likely to do any (total) MVPA in a typical week (AMEs boys: 3%; 95% CI: 0% to 6%; girls: 4%; 95% CI: 0% to 7%); whilst girls in high-income versus low-income households spend more time being active (AMEs total MVPA: 16 min/day, 95% CI: 5 to 26; total MVPA-active: 14 min/day, 95% CI: 3 to 25). As in England, the findings for total MVPA masked differences by gender, domain and outcome.

First, higher levels of recreational MVPA in high-income versus low-income households were evident among both genders and each outcome. For example, differences between adolescents in high-income versus low-income households in recreational MVPA were 15 min/day in a typical week (95% CI: 6 to 24) among boys and 19 min/day (95% CI: 12 to 27) among girls; differences in recreational MVPA-active were 12 min/day (95% CI: 2 to 21) and 16 min/day (95% CI: 8 to 24) for boys and girls, respectively.

Secondly, boys in high-income households were more likely to do any work-based MVPA than their counterparts in low-income households (AME 9%; 95% CI: 2% to 16%), yet the quantitative outcomes (MVPA and MVPA-active) showed similar levels by income. Thirdly, adolescents in low-income versus high-income households were more likely to have taken part in active transportation (AMEs boys: 11%; 95% CI: 3% to 19%; girls: 10%; 95% CI: 3% to 17%). Among all participants(including those who did none in a typical week), those in the lowest-income versus highest-income households spent 7 min/day more in active travel (AMEs travel MVPA boys: 95% CI: 3 to 12; girls: 95% CI: 4 to 9). Among those who did any, boys and girls in low-income versus high-income households spent 8 min/day (95% CI: 1 to 14) and 11 min/day (95% CI: 7 to 16) longer on average travelling actively in a typical week, respectively.

## DISCUSSION

Using nationally representative data from adolescents in England and the US, hurdle models were applied to compare levels and inequalities in self-reported total and domain-specific MVPA. We hypothesised that adolescents in high-income households were more likely both to participate in MVPA and, conditional on doing any, to spend more time on average being active than their counterparts in low-income households. Our analyses revealed a more complex picture: differences in MVPA by household income status varied by gender, domain, and outcome. Levels of formal sports/exercise and recreational MVPA were higher among adolescents in high-income households in England and the US, respectively. In contrast, levels of active travel, among boys in England and both genders in the US, were higher in low-income households.

### Comparisons with previous studies

Comparisons with previous studies are difficult due to differences in study characteristics (e.g. age range, or use of objective, device-based measurement) and analytical strategy. Bearing in mind this caveat, the low levels of MVPA across all income groups presented here agree with other English and US studies. In England, data from the HSE 2015 showed that 21% and 16% of boys and girls aged 5–15 years respectively achieved the WHO recommendation of at least 60 minutes of MVPA per day;[12] US data from the 2016 National Survey on Children’s Health (NSCH) showed an equivalent figure of 24% among participants aged 6–17 years.[29]

Likewise, the evidence of inequalities presented here broadly agree with systematic reviews,[30] findings for VPA among children in the UK,[31] and US findings for physical activity,[32] inactivity,[21] recreational activity,[20] active transportation [29] and cardiorespiratory fitness.[33] Our findings also agree with worldwide studies for levels of activity outside-of-school [25] and for activity frequency.[34]

Our findings relating to the domain-specific nature of inequalities are also in agreement with previous studies. The lower involvement of adolescents in low-income households in formal, structured sports/exercise activities corresponds with empirical studies in England [35] and Australia.[5] Our findings of divergent patterns in the recreational and active transportation domains in the US correspond with similar patterns found among adults using the same datasets as the present study.[36] Likewise, the higher levels of active travel for boys in low-income households in England agrees with findings of a greater likelihood of active travel among adults in more deprived areas in Scotland.[37] UK studies of younger children (7–8 year-olds) using accelerometry suggest no clear socioeconomic gradients in the time spent in MVPA;[38] however, activity monitors do not currently capture data on activity domain.

Our novel use of hurdle models adds to recent literature by showing the domain- and outcome-specific nature of inequalities in adolescent MVPA. For example, boys in England in high-income households were more likely to do any formal sports/exercise MVPA than their counterparts in low-income households; whilst the amount of time spent doing sports/exercise showed no difference by income amongst those who did any. In contrast, inequalities in recreational activity in the US exist in participation and in time spent being active. Such findings illustrate the limitations of using single equation regression models when the determinants for participation and duration may differ. Decomposing the single quantitative MVPA variable via hurdle modelling can therefore potentially shed light on the determinants of inequalities in the lower-tail of the distribution (drivers of inactivity) and those impacting the positive, non-zero, part of the distribution, implying potentially different solutions to reduce inequalities.[39]

### Mechanisms and implications for policy

There are numerous pathways through which markers of SEP such as household income impact on physical activity. Differences in financial/wealth resources and the built environment, including those driving inequalities in opportunity and access to affordable facilities and safe public outdoor spaces [40] are likely key modifiable determinants of inequalities in formal (England) and recreational (US) activities. Higher levels of active travel in low-income households likely reflect lower car ownership.[41] Improving overall levels of PA and reducing inequalities requires policy actions and interventions to ensure low barriers of entry and adequate support to enable adolescents to “move more and sit less”.[42] Tackling income-based inequalities would also require tackling disparities in PA by the correlated dimension of race/ethnicity.[43] Analyses of NHANES 2011–12 data show lower levels of adolescent MVPA among non-Hispanic Blacks, Hispanics and Asians compared with non-Hispanic White populations.[44]

According to the WHO,[45] reducing inequalities requires both population-based policy actions to tackle the “upstream” determinants that shape the equity of opportunities for PA and support for “downstream” individually-focused (educational and informational) interventions, with both implemented according to the principle of proportional universality. Examples of the former include encouraging non-motorised travel modes (through better road connectivity and improved provision of cycling and walking infrastructure such as segregated cycle lanes and improved road safety through traffic free routes[46]), and creating more opportunities for PA in public open spaces and local community settings.[3] In the UK, large PA interventions focusing on knowledge and motivation among primary school children have yielded null findings, highlighting the importance of more upstream approaches.[47]

### Strengths and limitations

Strengths of our study include the use of nationally representative data across PA domains. Although it is well-known that MVPA distributions typically contain excess zeros and positive skew, no epidemiological studies to date have applied hurdle models to assess the different aspects of adolescent MVPA (participation and duration) and estimate inequalities in these. Hurdle models avoid the loss of information and statistical power that occurs when the quantitative MVPA variable is grouped into a binary variable, transformed to meet the assumption of normality,[7] or when analysed in a single-equation model.[20]

Caution is required, however, when interpreting our findings. First, self-reported PA data has well-known limitations such as recall and reporting (social desirability) bias;[48] this may be socially patterned, thereby potentially upwardly or downwardly biasing our estimates of inequalities. Secondly, the analytical sample sizes (reduced further by missing income data) means our findings will be statistically underpowered to some extent despite the pooling of data across survey years. Thirdly, the analytical samples (aged 11–15 and 12–17 in England and the US, respectively) and PA outcomes were different. However, our aim was to compare inequalities rather than levels of MVPA. Fourthly, the choice of potential confounders was limited by data availability. We were unable to provide separate estimates by race/ethnicity using NHANES data or examine any potential moderation of income inequalities. Fifthly, our findings are contingent upon HSE and NHANES data collection methods, including the exclusion of in-school MVPA and the assumption that all activities were of at least moderate-intensity (HSE), the minimum duration of 10 minutes in NHANES (in accord with the contemporaneous US guidance [49] but differing from recent guidelines which acknowledge that PA of any duration enhances health[50]), and the inability to specifically focus on inequalities in PA that typically require financial resources (both datasets). We acknowledge that different definitions may have led to different conclusions. Finally, we cannot draw causal inferences, as this was a descriptive study based on cross-sectional data.

## CONCLUSIONS

Participation in formal sports/exercise and recreational MVPA was higher among adolescents in high-income households in England and the US, respectively. Our findings may assist policy-makers to identify and commission tailored policy actions and interventions to reduce inequalities, and our methods could be used by practitioners to monitor and evaluate their impact.

## Data Availability

HSE datasets are available via the UK Data Service (http://www.ukdataservice.ac.uk). NHANES datasets are available via the CDC website (https://www.cdc.gov/nchs/nhanes).

## Acknowledgements

The authors thank the interviewers and nurses, the participants in the Health Survey for England series, and colleagues at NatCen Social Research.

## Author contributions

SS conceptualised the study. SS was responsible for conducting the analyses, interpreting the results and drafting the manuscript. SS and JSM critically revised the manuscript. Both authors have read and approved the final manuscript.

## Patient consent

Not required.

## Ethics approval

Research ethics approval for the HSE 2008 and HSE 2012 was obtained from the Oxford A Research Ethics Committee (reference number 07/H0604/102 and 10/H0604/56, respectively); ethical approval for the HSE 2015 was obtained from the West London Research Ethics Committee (reference number 14/LO/0862). NHANES protocols were approved by the National Center for Health Statistics Ethics Review Board.

## Strengths and limitations of this study

- In contrast to single-equation regression modelling, hurdle models are well suited to analysing quantitative variables such as time spent in moderate-to-vigorous physical activity (MVPA) as separate models are estimated for participation and the amount of time spent active(conditional on overcoming the “hurdle” of participation).
- This study applies hurdle models to nationally representative data from England and the United States to estimate inequalities in both aspects of MVPA among adolescents.
- Self-reported data on physical activity may contain recall and reporting (social desirability) bias.
- Causal inferences cannot be drawn, as this was a descriptive study based on cross-sectional data.

